# SUPPORT NEEDS AND ADAPTIVE BEHAVIOR SURVEYS: SERVICES PREDICTION AND RELATIONSHIP

**DOI:** 10.1101/2023.11.05.23298117

**Authors:** Annalisa V. Piccorelli, Eric J. Moody, David Heath, Ethan Dahl, Sandy Root-Elledge

**Affiliations:** Division of Biostatistics, Kaiser Permanente Washington Health Research Institute; Wyoming Institute for Disabilities, University of Wyoming

**Keywords:** Supports Intensity Scale, Inventory for Client and Agency Planning, Medicaid eligibility, disabilities

## Abstract

The Inventory for Client and Agency Planning (ICAP) and the Supports Intensity Scale (SIS) have been used to determine Medicaid eligibility for individuals with disabilities but are designed to capture different information. This project explored how these surveys relate to services received and each other. ICAP and SIS surveys were conducted on 125 [Blinded] adults with intellectual and developmental disabilities eligible for a Medicaid Waiver. Results suggest that both measures were strongly associated and could be used to predict services. However, this study suggests that the SIS and ICAP summary measures were separate constructs. Therefore, both instruments can be used to determine Medicaid eligibility, but implementers should be aware of differences in the types of constructs being captured.

## Introduction

The Home and Community Based Services (HCBS) Medicaid waivers allows states to fund disability services delivered in the community (Braddock et al., 2005; Moseley, 2005). Individual support budgets are allocated based on a determination of the clients’ need and aligning the funding level to their service requirements (Arnold et al., 2015; Moseley, 2005). The method used to determine needs and allocate funding varies from state to state (Moseley, 2005). Some states developed their own tool, but the majority of states, (and some Canadian provinces), use the Inventory for Client and Agency Planning (ICAP; Bruininks et al., 1986a) and/or the Supports Intensity Scale (SIS; Thompson et al., 2004a) in their process (American Association of Intellectual and Developmental Disabilities, 2020; Fortune et al., 2008; Weiss et al., 2009).

Both tools have been extensively studied and have good psychometric properties (Bruininks et al., 1986b; Buntinx et al., 2008; Chou et al., 2013; Jenaro et al., 2011; Thompson et al., 2004a, 2004b; Thompson et al., 2002; Thompson et al., 2008). However, there is little information on how these tools relate to individual funding levels and whether these tools capture similar information. Indeed, both tools were designed quite differently. The ICAP is older and interviews clients to determine their adaptive and maladaptive behaviors, focusing on what they *can and cannot do independently* (Bruininks et al., 1986a). The ICAP assesses adaptive behavior on the basis of motor, social and communication, personal and community skills; the skills needed by individuals with intellectual and developmental disabilities (IDD) to participate in their daily lives (Bruininks et al., 1986a; Luckasson et al., 2002). Maladaptive behavior is assessed according to frequency and severity of problem behaviors, such as self-injurious behavior, disruptive behavior, and social withdrawal (Bruininks et al., 1986a). Funding allocation algorithms among the states are diverse in how they use the ICAP. For example, Wyoming uses the ICAP summary measure, service level, the maladaptive behavior section along with age and living situation; Illinois requires the ICAP in addition to psychological and medical assessments (Illinois Department of Human Services, 2019; Wyoming Department of Health, 2014). The SIS, by contrast, is newer and focuses on the supports individuals require to participate in their community, *regardless of any underlying condition or disability* (Brown et al., 2009; Guscia et al., 2006; Harries et al., 2005; Thompson et al., 2002; Thompson et al., 2009; Wehmeyer et al., 2009).

Although many states continue to use the ICAP to determine HCBS funding levels, there are growing calls for the SIS to be included in the process based on the positive relationship between funding and support needs (Chou et al., 2013). For instance, the number of support needs is associated with the need for greater funding (Fortune et al., 2008) suggesting that there may be no need to assess maladaptive behaviors at all (Arnold et al., 2015). However, like the ICAP, the SIS may be used in conjunction with other tools. For instance, in Louisiana, the SIS is used with the Louisiana Plus, which assesses a variety of specific supports needs and risk factors to determine funding allocation (Louisiana Department of Health and Hospitals, 2012). Tennessee conducts follow-up interviews after the SIS to determine additional details regarding extraordinary needs (Tennessee Department of Intellectual and Developmental Disabilities, 2019). Moreover, the ICAP and SIS can be used together (e.g., North Dakota). For example, adaptive behavior (assessed by the ICAP) and support needs (assessed by the SIS) both explain an individual’s way of participating in daily life (Thompson et al., 2002). The SIS does not measure the ability of a person to carry out everyday activities like the ICAP (Weiss et al., 2009).

Importantly, several studies have demonstrated a significant correlation between the SIS and ICAP summary scores (Guscia et al., 2006; Harries et al., 2005; Thompson et al., 2002), but there is disagreement on whether the ICAP and SIS are measuring the same constructs. Thompson et al. (2002) suggests that these tools describe different aspects of an individual’s way of participating in daily life. However, Harries et al. (2005) demonstrated the ICAP, SIS, along with the Adaptive Behavior Scale-Residential and Community (ABS-RC: 2) measure the same construct using principal components analysis. No studies have attempted to resolve the relationship between SIS and the ICAP and their association with funding of individuals with IDD. Thus, this study explores the relationship between the ICAP and the SIS, and their relation to funding. We used the types of services a client receives (residential facility or school/day program) as a proxy for individual funding level. Thus, the primary aim of this study was to determine the association between the ICAP and the SIS and services, as the driver of individual budget. The secondary aim of this study is to examine the associations between these two instruments to determine if they are capturing similar constructs and compare our results to prior studies.

## Methods

### Participants

This study was approved the [Blinded] Institutional Review Board. The study participants consisted of 125 [Blinded] adults with IDD who qualify for an HCBS Waiver in 2015. Participants were eligible if they were due to have an ICAP scheduled sometime during the year that the study took place. These ICAPs were all being collected as part of standard HSBC procedures for the state to update their funding levels. A sample of eligible clients was randomly selected for the SIS assessment by the [Blinded] to ensure raters could not impact the sample. All data for this analysis came from a program that conducts level of care assessments for Medicaid Waiver participants. The ICAP was administered to determine level of care for programmatic purposes. The SIS was administered as part of a study commissioned by the [Blinded] Department of Health to assess the viability of using the SIS for level of care determination. As part of this ICAP program, written consent was obtained from the participant, their parents or guardians. Separate written consent was obtained from participants, caregivers or guardians for the SIS given that it was not being used for programmatic purposes, but to explore its use by the program. Given that this analysis is a secondary data analysis of programmatic data, all data use in this analysis were deidentified, and then entered into an analytic database by program staff before being provided to the researchers. [Blinded] IRB granted an exemption for this analysis because all data were deidentified.

From January 1 to December 31, 2015, all eligible individuals were invited to participate in this study at the time of their scheduled ICAP until the desired sample size was achieved. Currently, the ICAP is used to determine the Level of Service Need (LSN) in [Blinded] ([Blinded]). The LSN, age group, and living situation are used to determine Individual Budget Amount ([Blinded]). See Table 1 for demographics.

**Table 1.**
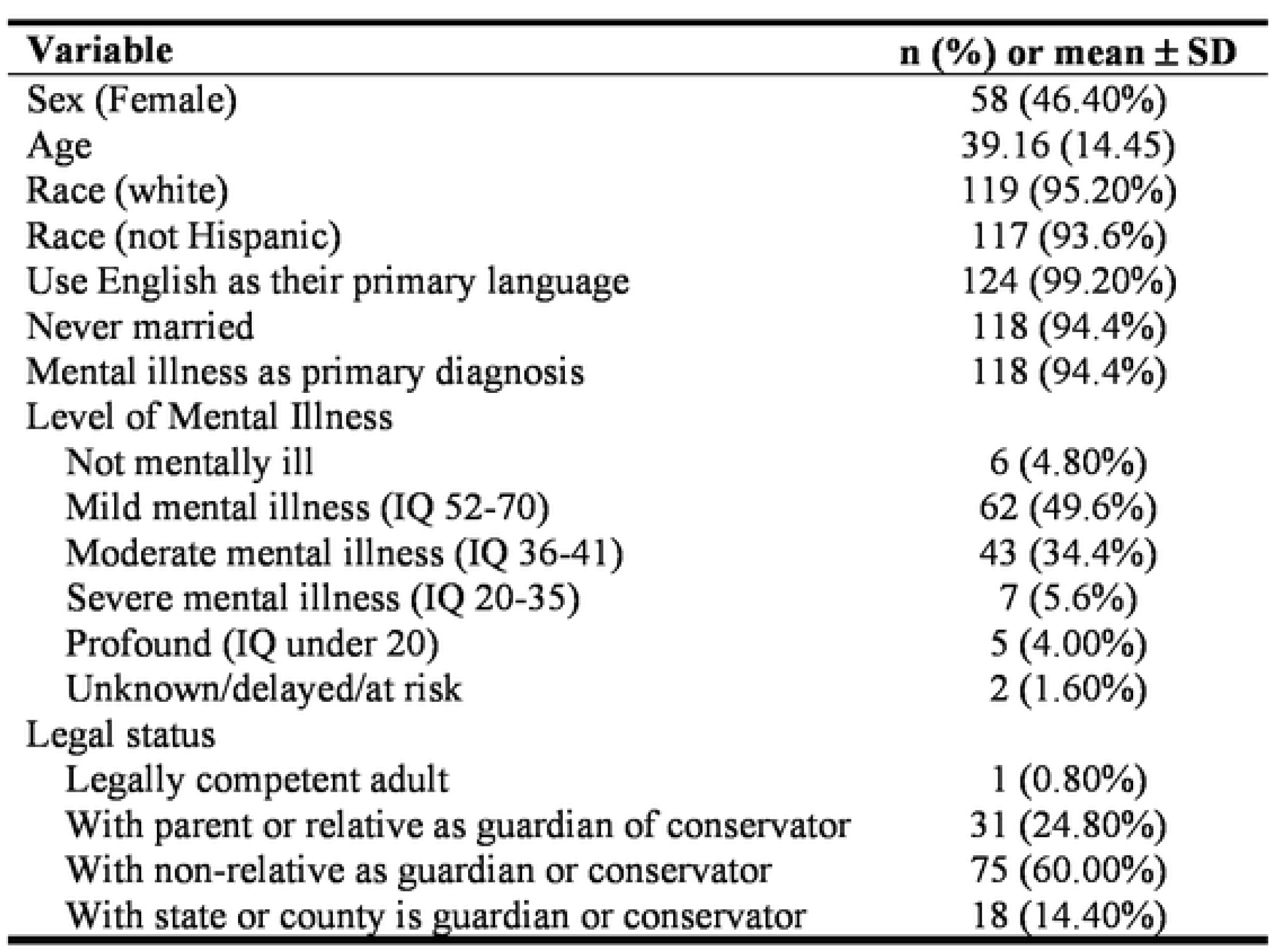
Sample Demographics.

### Measures

#### Supports Intensity Scale (SIS)

The SIS determines the frequency and intensity of support needs using the Support Needs Scale, which is made up of six subscales of activities of everyday life, and Supplemental Protection and Advocacy activities (Thompson et al., 2004a, 2004b). The SIS also includes a description of the Exceptional Medical and Behavioral Support Needs (Thompson et al., 2004a, 2004b). The SIS is composed of the following three sections: Section 1. Support Needs Scale, Section 2. Supplemental Protection and Advocacy Scale, and Section 3. Exceptional Medical and Behavioral Support Needs (Thompson et al., 2004a). This study focused on the responses to the SIS Section 1, which describes the frequency and intensity of supports needed to carry out life activities. Section 1 includes the following 6 subscales: (a) Home Living Activities, (b) Community Living Activities, (c) Lifelong Learning Activities, (d) Employment Activities, (e) Health and Safety Activities, and (f) Social Activities (Thompson et al., 2004a). The life activities are rated on 5-point scales for frequency of how often the support is needed, from *none or less than monthly* (0) to *hourly or more frequently* (4), duration of time required to provide the support each day, from *none* (0) to *4 hours or more* (4), and type of support, from *none* (0) to *full physical assistance* (4) for each item (Thompson et al., 2004a). Section 1 is summarized with the SIS Supports Needs Index (SIS SNI), which takes the raw scores from the Activities subscales into account, and the SIS SNI Percentile (Thompson et al., 2004b). The psychometric properties of the SIS have demonstrated high internal consistency (0.94-0.99), interrater (0.55-0.90) and test-retest (0.74-0.94) reliability, content, criterion and construct validity, and discriminating power (0.66-0.72) (Buntinx et al., 2008; Chou et al., 2013; Jenaro et al., 2011; Thompson et al., 2004b; Thompson et al., 2002; Thompson et al., 2008). The six-subscale structure of the Support Needs Scale was confirmed by a factor analysis (Kuppens et al., 2010). Supplemental Protection and Advocacy activities and Exceptional Medical and Behavioral Support Needs have been demonstrated to strongly associate with the frequency and intensity of the support needs of the Support Needs Scale of the SIS in a sample of adolescents aged 16-22 years (Seo et al., 2017).

#### Inventory for Client and Agency Planning (ICAP)

The ICAP is comprised of the following ten sections: Section A. Descriptive Information, Section B. Diagnostic Status, Section C. Functional Limitations and Needed Assistance, Section D. Adaptive Behavior, Section E. Problem Behavior, Section F. Residential Placement, Section G. Daytime Program, Section H. Support Services, Section I. Social and Leisure Activities, and Section J. General Information Recommendations (Bruininks et al., 1986a). In addition to these is a maladaptive behavior worksheet. For this study, Section D. Adaptive Behavior is of particular interest. It is made up of the following subsections: (a) Motor Skills, (b) Social and Communication Skills, (c) Personal Living Skills, and (d) Community Living Skills (Bruininks et al., 1986a). The score for each skill is recorded based on a 4-point scale from *never or rarely* (0) to *does very well* (4) in response to “Does (or could do) task completely without help or supervision” (Bruininks et al., 1986a). The respondent is thinking in terms of what the client can do (Bruininks et al., 1986a). The summary measure of Section D. Adaptive Behavior is the Total Adaptive Behavior Raw Score (Bruininks et al., 1986a). Section E. Problem Behavior measures the frequency and severity of the following problem behaviors: (a) Hurtful to Self, (b) Hurtful to Others, (c) Destructive to Property, (d) Disruptive Behavior, (e) Unusual or Repetitive Habits, (f) Socially Offensive Behavior, (g) Withdrawal or Inattentive Behavior, and (h) Uncooperative Behavior (Bruininks et al., 1986a). The ICAP Service Level is a composite of the Total Adaptive Behavior Raw Score with the General Maladaptive Index (calculated based on Section E. Problem Behavior) and meant to describe the relationship between these behaviors and services required (Bruininks et al., 1986b). The ICAP has high internal consistency (0.85 to 0.88), test-retest (0.74 to 0.88) and interrater reliability (67% to 100%), and construct, content, and criterion-related (0.91 to 0.99) validity (Bruininks et al., 1986b).

Based on the responses to Residential Facility and School/Day Program within ICAP Section A. Descriptive Information, a variable describing services, labeled SERVICES, was created. To create SERVICES, Residential Facility responses were divided into the following three categories: 1) Family, 2) Supported Living and 3) Residential Habilitation home services. The latter two categories made up the first two categories of the SERVICES variable. The Family category was combined with the School/Day Program to create the final two categories of SERVICES because those who received Day Habilitation services were categorized as Day Supported Living and those who were employed, in school, or had none for School/Day Program were categorized as Other. Therefore, the SERVICES variable was made up of the following four categories: Other (*n* = 17, 13.6%), Supported Living (*n* = 17, 13.6%), Day Supported Living (*n* = 32, 25.6%), and Residential Habilitation (*n* = 59, 47.2%). Those who fell in the Other category were assumed to receive minimal Medicaid services and were expected to have the lowest funding. This study set up the order of the SERVICES categories based on order of funding from lowest to highest as follows: Other = 1, Supported Living = 2, Day Supported Living = 3, and Residential Habilitation = 4. This is based on the assumption that Day Supported Living Services are more intense and time consuming than Supported Living Services and thus, cost more.

### Procedure

Respondents were individuals who have known the client for at least 3 months and were interviewed in person. Respondents included the client’s parents/guardians, teachers, providers, nurses, vocational rehabilitation counselors, and mental health counselors. For the ICAP, the raters met with at least two of the respondents independently or separately. For the SIS, the raters met with the respondents in a team setting, with the individual with IDD joining the meeting roughly 25% of the time.

The raters had experience working as case managers or day service providers as well as extensive experience (34 combined years) doing the ICAP assessments. The raters were trained on the ICAP as part of their work duties and maintained fidelity throughout the study. For the SIS, the raters were trained and certified by the American Association on Intellectual and Developmental Disabilities. Raters were familiar with the respondents because of working with them to complete ICAPs for several years. The ICAP was completed prior to the SIS in all cases.

### Analysis Plan

The first aim of this study was to determine the association between SIS outcomes and current service level (SERVICES). SERVICES was the dependent variable in this study. The state uses the ICAP summary measures along with other details about the participant to compute the Individual Budget Amount ([Blinded]). Thus, the ICAP was highly associated with the services received in this dataset. As a result, in the models predicting SERVICES, the summary measures of the SIS were the independent variables, controlling for ICAP by including the ICAP summary measure (ICAP Service Level) as a covariate. The secondary aim of this study was to examine the association between the ICAP and the SIS to explore if they capture similar constructs and compare our results to prior studies. To carry out this aim, we analyzed correlations and conducted principal components analysis.

Analyses were conducted using SAS® 9.4 (*SAS*, 2018). One-way Analysis of Variance (ANOVA) was used to determine the association between the summary measures of the SIS and the ICAP, the SIS SNI and Percentile, and the ICAP Service Level, respectively, and SERVICES (Other = 1, Supported Living = 2, Day Supported Living = 3, and Residential Habilitation = 4) to address Aim 1.

Next ordinal logistic regression was used to determine if the summary measures of the SIS, the SIS SNI and Percentile could predict SERVICES. Given the potential relationship between the ICAP Service Level and SERVICES, the ICAP Service Level was adjusted for in these models by its inclusion as a covariate. Variance inflation factor (VIF) greater than 10 was used to indicate multicollinearity. VIF > 10 were observed among the SIS SNI and the SIS SNI Percentile so they were not included in the same models. The Score Test for Proportional Odds Assumption determined the proportional odds assumption related to the order of the response variable was not met. However, the order of SERVICES made sense conceptually and allowed for the clearer interpretation and the estimated coefficients of the intercepts agreed with the order suggested. SERVICES = 1 (Other) was set as the baseline category for the ordinal logistic regression models. Odds ratios indicating the odds of an increase in SERVICES per one unit increase in the predictor were calculated.

This study also investigated the 6 subscales for the SIS Section 1. Support Needs Scale and the 4 subscales of Section D: Adaptive Behavior of the ICAP as predictors of SERVICES with an ordered logistic regression. VIF > 10 were observed among the SIS summary measures and subscales. The VIF of the ICAP Service Level was 9.84 when included with the ICAP subscales in a model. Since this approached our boundary of VIF > 10, ICAP Service Level and subscales were not included in the same model. Additionally, the ICAP subscales are components used to determine ICAP service level, thus, they should not be included in the same model. Stepwise ordered logistic regression was used to determine whether to include the subscales as predictors in the final model based on significance level. Given the larger number of predictors relative to sample size, the *p*-value boundary of entry of a predictor was ≤ 0.20 and the *p*-value boundary for removal of a predictor was > 0.20.

Spearman Rank-Order correlation was used to assess the association between the 6 subscales of the SIS and the 4 subscales of the ICAP and their respective summary measures, the SIS SNI and Percentile, and the ICAP Service Level. Spearman correlation coefficients were used because these variables were not normally distributed based on the Shapiro-Wilk Test of Normality.

Principal components analysis (PCA) of the 6 subscales for the SIS and the 4 sections of Section D. Adaptive Behavior of the ICAP was conducted. The following criteria were used to determine whether a factor should be included in the solution: first, eigenvalue > 1, and second, at or prior to the “elbow” in the scree plot (Johnson & Wichern, 2002; Kaiser, 1960; Kuhlfeld et al., 2010). When greater than one factor was indicated, an orthogonal varimax prerotation was completed followed by an oblique Procrustes rotation, which results in uncorrelated factors to facilitate interpretation (Jolliffe, 2002; Kuhlfeld et al., 2010). Interpretation of factors was based on absolute value of pattern coefficients greater than or equal to 0.5, indicating strong correlation between the subscale and the factor (Yung et al., 2010).

## Results

### Relationship Between SERVICES and the SIS Controlling for the ICAP

The results of the ANOVA are shown in Table 2. The ICAP and the SIS summary measures, all the ICAP subsections, and four of the SIS subscales (Home Living Activities, Community Living Activities, Employment Activities, and Health and Safety Activities) were found to be significantly associated with SERVICES (all *p* ≤ 0.05). The SIS subscale, Life-Long Learning Activities, was moderately associated with SERVICES, while the SIS subscale, Social Activities, did not have evidence of an association with SERVICES. The mean ICAP Service Level for Supported Living and Other were in the range 60-69 described as between “Regular personal care and/or close supervision” and “Limited personal care and/or regular supervision” (Bruininks et al., 1986a). For Residential Habilitation and Day Supported Living, the mean ICAP Service was in the range 50-59 described as “Regular personal care and/or close supervision” (Bruininks et al., 1986a). Higher SIS SNI and SIS SNI Percentile indicates greater need of supports (Thompson et al., 2004a). The mean SIS SNI and mean SIS SNI Percentile suggested amount of support needs for those with Residential Habilitation and Day Supported Living is more than those that Supported Living and Other SERVICES.

**Table 2.**
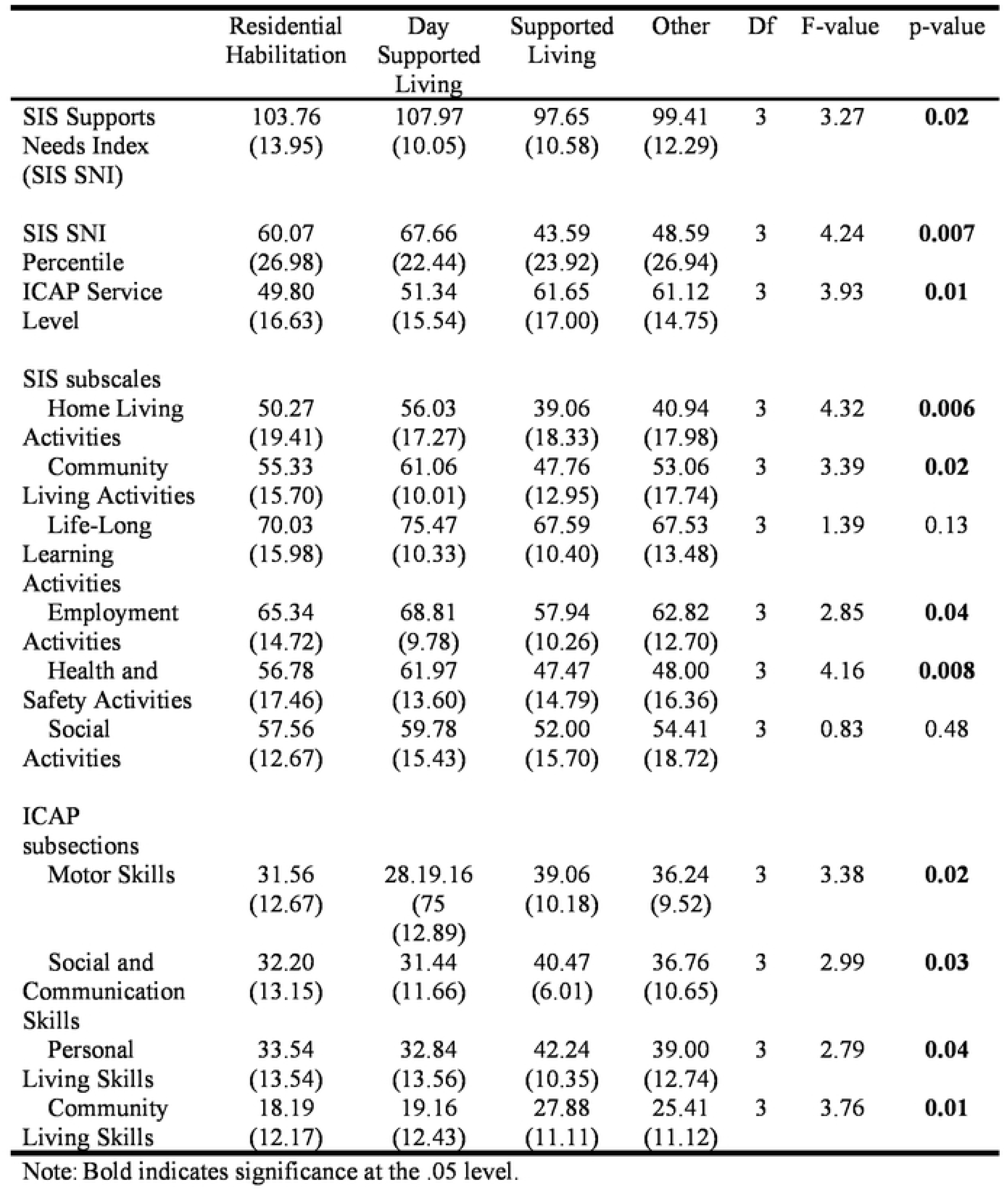
Means and Standard Deviations of the SIS and ICAP Summary and Subscale Measures included in SERVICES, and F-tests of Association between the Summary Measures and SERVICES.

The ordinal logistic regression results are shown in Table 3. As expected, ICAP Service Level proved to be the major indicator of SERVICES in the simple model and the multivariate models including the SIS summary measures, *χ^2^* (1) = 8.42, *p* = 0.004, 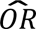= 0.97, 95% CI = 0.95, 0.99; *χ^2^* (1) = 7.66, *p* = 0.006, 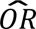= 0.96, 95% CI = 0.93, 0.99; and *χ^2^* (1) = 5.64, p = 0.02, 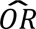= 0.97, 95% CI = 0.94, 0.99, respectively. The estimated odds ratio of 0.97 for the simple model can be interpreted as the estimated odds of an increase in the level of SERVICES decrease by 3.0% per 1 unit increase in ICAP Service Level. In the multivariate models, there was not evidence that the SIS SNI and Percentile had an effect on SERVICES while controlling for ICAP Service Level, *χ^2^* (1) = 0.90, *p* = 0.34, 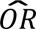= 0.98, 95% CI = 0.95, 1.02; and *χ^2^* (1) = 0.17, *p* = 0.68, 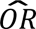= 1.00, 95% CI = 0.98, 1.01, respectively.

**Table 3.**
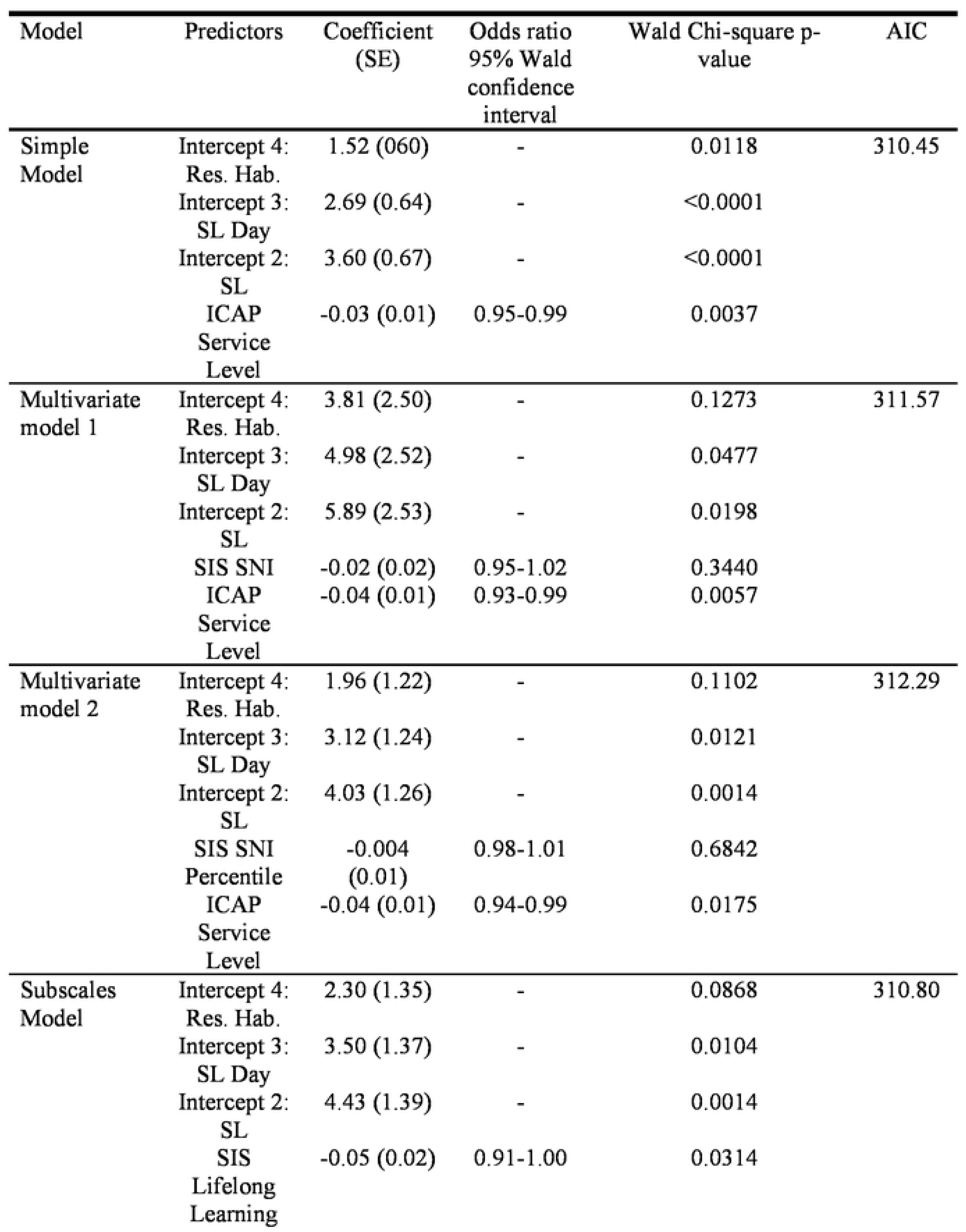
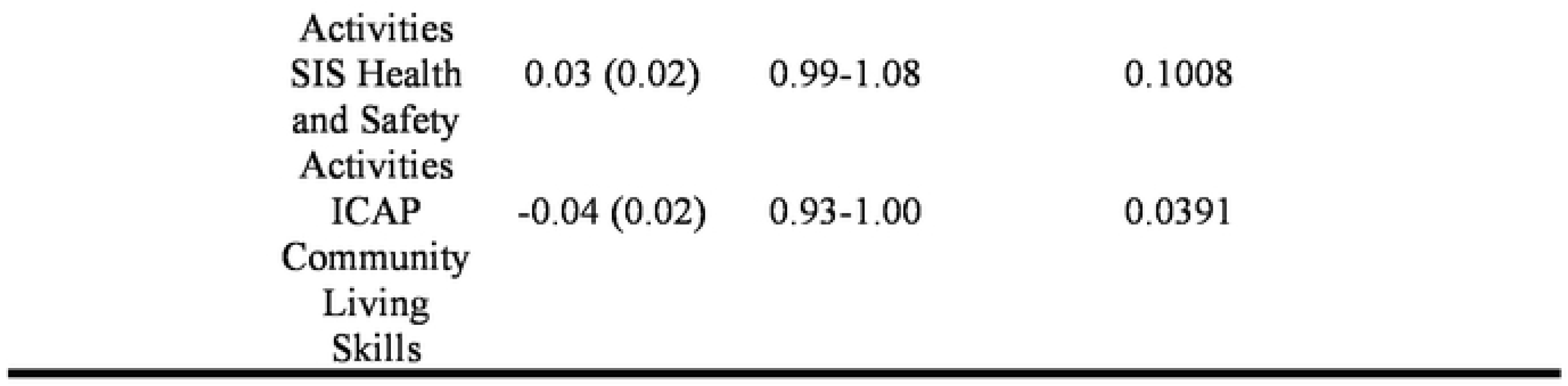
Ordinal Logistic Regression Models of SERVICES Using SIS and ICAP.

The ICAP Community Living Skills and SIS Lifelong Learning and SIS Health and Safety Activities subscales were selected in the Subscales Model. SIS Lifelong Learning Activities and ICAP Community Living Skills were significant predictors (p < 0.05) of SERVICES, *χ^2^* (1) = 4.63, p = 0.03, 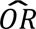= 0.95, 95% CI = 0.91, 1.00, and *χ^2^* (1) = 4.62, p = 0.04, 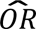= 0.96, 95% CI = 0.93, 1.00, respectively. There was moderate evidence that SIS Health and Safety Activities had an effect on SERVICES, *χ^2^* (1) = 2.69, p = 0.1, 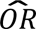= 1.04, 95% CI = 0.99, 1.08. The estimated odds of an increase in the level of SERVICES decrease by 4.9% per 1 unit increase in SIS Lifelong Learning Activities, while controlling for ICAP Community Living Skills and SIS Health and Safety Activities. The estimated odds of an increase in the level of SERVICES decrease by 3.7% per 1 unit increase in ICAP Community Living Skills, while controlling for SIS Community Living and SIS Health and Safety Activities.

### Relationship between the SIS and the ICAP

The summary measures of the SIS Section 1 Supports Needs Scale and the ICAP Section D Adaptive Behavior are shown on Table 4. The mean ICAP Service Level of 53.54 falls in category of “Regular personal care” (range: 50-59) and the range of 9 to 86 almost encompasses the full range of the amount of care as described by the ICAP, from “Total personal care and intense supervision” to “Infrequent or no assistance for daily living” (Bruininks et al., 1986a). The Support Needs Index (SNI) ranging from 54 to 124 and the SIS SNI Percentile ranging from 1 to 95 indicate the sample encompasses the full range of support needs from those that have low intensity and infrequent support needs to those with high intensity and frequent support needs.

**Table 4.** Descriptive Measures of the SIS Support Needs Scale and the ICAP Adaptive Behavior and Service Level.

| Measure | Mean (SD) | Median | Range |
| --- | --- | --- | --- |
| SIS Section 1 Support Needs Scale |  |  |  |
| Home Living Activities | 48.95 (19.28) | 50.00 | 0-79 |
| Community Living Activities | 55.46 (14.82) | 60.00 | 0-84 |
| Life-Long Learning Activities | 70.75 (13.87) | 70.75 | 0-94 |
| Employment Activities | 64.88 (13.07) | 68.00 | 0-87 |
| Health and Safety Activities | 55.78 (16.66) | 57.00 | 0-83 |
| Social Activities | 56.94 (17.90) | 59.00 | 0-84 |
| SIS Supports Needs Index (SIS SNI) | 103.42 (12.74) | 106.00 | 54-124 |
| SIS SNI Percentile | 58.21 (26.47) | 65.00 | 1-95 |
| ICAP Section D. Adaptive Behavior |  |  |  |
| Motor Skills | 32.50 (12.39) | 36.00 | 0-51 |
| Social and Communication Skills | 33.75 (12.01) | 37.00 | 2-52 |
| Personal Living Skills | 35.29 (13.34) | 39.00 | 1-58 |
| Community Living Skills | 20.74 (12.39) | 21.00 | 0-45 |
| ICAP Service Level | 53.34 (16.73) | 56.00 | 9-86 |

Note: Medians are also reported because the summary measures were not normally distributed based on the Shapiro-Wilk Test of Normality.

### Correlations

To understand the relationship between the SIS and the ICAP, the Spearman correlation coefficients between the SIS Support Needs Scale subscales and the ICAP Adaptive Behavior subsections and the summary measures of the SIS and the ICAP, the SIS SNI and SNI Percentile and the ICAP Service Level, respectively, were investigated (Table 5). The correlations were all negative as expected based on the opposing set up of the ICAP compared to the SIS, e.g., a “3” indicates “DOES VERY WELL” and “0” indicates “NEVER OR RARELY” in terms of a subject completing a task without help for the ICAP whereas for the SIS a “3” indicates “at least once of day” and “0” indicates “none of less than monthly” in terms of how often a subject needs help with a given activity (Bruininks et al., 1986a; Thompson et al., 2004a). The correlations ranged from *r* = -0.44 to *r* = -0.76. Strong correlation coefficients (|*r*| ≥ 0.5) ranged from -0.72 to -0.50 and were observed in the majority of the associations between the subscales. The summary measures of the SIS and the ICAP were strongly correlated at -0.75. All correlation coefficients were significant at *p* <0.0001.

**Table 5.** Spearman correlation coefficients between the SIS subscales and ICAP Adaptive Behavior subsections.

| Subsection | ICAP |  |  |  |  |
| --- | --- | --- | --- | --- | --- |
|  | Motor Skills | Social and Communication Skills | Personal Living Skills | Community Living Skills | ICAP Service Level |
| SIS |  |  |  |  |  |
| Home Living Activities | -0.66 | -0.71 | -0.75 | -0.73 | -0.76 |
| Community Living Activities | -0.51 | -0.64 | -0.57 | -0.63 | -0.65 |
| Life-Long Learning Activities | -0.49 | -0.62 | -0.56 | -0.64 | -0.62 |
| Employment Activities | -0.47 | -0.62 | -0.55 | -0.65 | -0.60 |
| Health and Safety Activities | -0.64 | -0.67 | -0.68 | -0.71 | -0.71 |
| Social Activities | -0.44 | -0.60 | -0.51 | -0.59 | -0.63 |
| SIS Supports Needs Index (SIS SNI) | -0.61 | -0.72 | -0.69 | -0.74 | -0.75 |
| SIS SNI Percentile | -0.61 | -0.72 | -0.69 | -0.74 | -0.75 |

Note: All correlation coefficients (r) were significant at the <0.0001 level.

### Principal Components Analysis

To further describe the relationship between the SIS and ICAP, principal components were identified by PCA of the subscales of the SIS Support Needs Scale and the ICAP Adaptive Behavior (Tables 6 and 7). The first three factors explained 87.90% of the variation. The eigenvalues of these three factors prior to rotation were as follows: Factor 1: 6.75 (explaining 67.53% of the variation), Factor 2: 1.63 (explaining 16.26% of the variation), and Factor 3: 0.41 (explaining 4.11% of the variation). A two-factor solution was indicated since factors 1 and 2 had eigenvalues greater than 1 prior to rotation (Kaiser, 1960). The third factor fell in the elbow of the scree plot, indicating it may help further explain the total sample variance (Johnson & Wichern, 2002). Thus, a three-factor solution was also investigated. As described in the Methods section, since there was more than one factor in these solutions, an orthogonal varimax prerotation was completed followed by an oblique Procrustes rotation for both.

**Table 6.** Factor pattern matrix for the Two-factor Solution obtained from the Principal Components Analysis.

| Subscale | Commonality ($h^2$) | Factor 1 | Factor 2 |
| --- | --- | --- | --- |
| SIS Support Needs Scale |  |  |  |
| Home Living Activities | 0.85 | **0.74** | **-0.55** |
| Community Living Activities | 0.80 | **0.86** | -0.27 |
| Life-Long Learning Activities | 0.87 | **0.91** | -0.21 |
| Employment Activities | 0.80 | **0.87** | -0.20 |
| Health and Safety Activities | 0.89 | **0.83** | -0.44 |
| Social Activities | 0.83 | **0.88** | -0.23 |
| ICAP Adaptive Behavior Subscale |  |  |  |
| Motor Skills | 0.81 | -0.16 | **0.88** |
| Social and Communication Skills | 0.83 | -0.32 | **0.85** |
| Personal Living Skills | 0.91 | -0.24 | **0.93** |
| Community Living Skills | 0.78 | -0.39 | **0.79** |

Note: Note: Bold indicates $\geq 0.5$

**Table 7.** Factor pattern matrix for the Three-factor PCJ Solution.

| Subscale | Commonality ($h^2$) | Factor 1 | Factor 2 | Factor 3 |
| --- | --- | --- | --- | --- |
| SIS Support Needs Scale |  |  |  |  |
| Home Living Activities | 0.86 | **0.74** | -0.46 | -0.31 |
| Community Living Activities | 0.80 | **0.85** | -0.18 | -0.21 |
| Life-Long Learning Activities | 0.88 | **0.91** | -0.16 | -0.14 |
| Employment Activities | 0.81 | **0.87** | -0.10 | -0.21 |
| Health and Safety Activities | 0.90 | **0.84** | -0.38 | -0.24 |
| Social Activities | 0.83 | **0.87** | -0.09 | -0.27 |
| ICAP Adaptive Behavior Subscale |  |  |  |  |
| Motor Skills | 0.94 | -0.18 | **0.91** | 0.28 |
| Social and Communication Skills | 0.91 | -0.29 | 0.46 | **0.78** |
| Personal Living Skills | 0.93 | -0.24 | **0.78** | **0.51** |
| Community Living Skills | 0.92 | -0.35 | 0.36 | **0.81** |

Note: Bold indicates $\geq 0.5$

### Two-factor solution

The eigenvalues after rotation for the factors 1 and 2 of the two-factor solution were 4.68 and 3.70, explaining 46.81% and 36.98% of the variance, respectively. The pattern matrix for the two-factor solution is shown on Table 6. The communalities (*h*^2^) ranged from 0.78 to 0.91 with median 0.83. Factor 1 included all six subscales of the SIS and none of the ICAP. Thus, factor 1 appears to describe the concept of supports needed for home and community living, learning, employment, health & safety and social activities. Factor 2 included all four subscales of the ICAP as well as the Home Living Activities subscale for the SIS. Factor 2 explains the concept of adaptive behavior. This makes sense given the strong correlations between the SIS Home Living Activities subscale and the four ICAP subscales ranging from *r* = -0.66 to *r* = -0.76 (Table 5). The ICAP Motor Skills and Personal Living Skills subscales encompass many of the skills required for the SIS Home Living Activities subscale. The SIS Home Living Activities subscale was strongly correlated with all 4 of the ICAP subscales (see Table 4).

### Three-factor solution

The three-factor solution indicated by the scree plot identified the following eigenvalues for factors 1, 2, and 3 after rotation: 4.61 (accounting for 46% of the variance), 2.23 (accounting for 22% of the variance) and 1.95 (accounting for 19.49%). The communalities were slightly higher for the three-factor solution, ranging from 0.80 to 0.94. Factor 1 encompassed all six subscales of the SIS, describing the concept of support needs. The ICAP subsections Motor and Personal Living Skills were included in Factor 2. The correlation between these subscales was strong at 0.77. Factor 3 described ICAP subsections Social and Communication, Personal and Community Living Skills. The correlations between these were strong, ranging from *r* = 0.79 to *r* = 0.88.

## Discussion

This study demonstrates a relationship between service level (SERVICES) and the SIS and ICAP summary measures. Interestingly, the mean SIS SNI, SIS SNI Percentile, and ICAP Service Level were similar between the Supported Living and Other, and between Residential Habilitation and Day Supported Living. ICAP Service Level was a significant predictor of SERVICES in simple and multivariate models. ICAP Community Living Skills subscale, and SIS Lifelong Learning Activities subscale were significant predictors of SERVICES in a multivariate model including the SIS Health and Safety Activities subscale. The SIS Health and Safety Activities subscale was also moderately significant in predicting SERVICES used by individuals with IDD, while controlling for ICAP Service Level. Given SERVICES is an indicator of funding, this study suggests both the SIS and ICAP could be used to predict or derive funding allocation. Therefore, both instruments can be used by state agencies to determine Medicaid funding.

However, we also found evidence that these instruments capture different constructs. We found strong associations between the summary measures and subscales of the SIS and the ICAP. Although the SIS and the ICAP are strongly associated, the 2-factor PCA resulted in the first factor solely encompassing supports needs as described by the 6 subscales of the SIS and the second factor including the 4 subsections of the ICAP and 1 subscale of the SIS, SIS Home Living Activities subscale. The 3-factor solution supported this as well with the first factor including only the 6 subscales of the SIS. The second and third factors included the ICAP subsections, Motor and Personal Living Skills, and the ICAP subsections, Social and Communication, Personal and Community Living Skills, respectively. The ICAP Motor and Personal Living Skills subsection making up the second factor can be attributed to the practical relationship between these subsections: motor skills are need to carry out personal skills, e.g., “picks up small objects with one hand” is necessary to “picks up and eats foods such as crackers” (Bruininks et al., 1986a). The association between the ICAP subsections, Social and Communication, Personal and Community Living Skills subsections in the third factor can be explained as these skills are what are needed to participate in social and personal life. Therefore, agencies using these tools to determine funding should be aware that they capture distinct information.

The correlation and PCA results were similar to those found by Thompson et al. (2002) but differed from the study by Harries et al. (2005). A number of factors could explain the differences in the PCA results between the current manuscript and Harries et al. (2005). Harries et al. (2005) included the adaptive behavior factors of the ABS-RC: 2 in their analysis, which could result in the different principal components identified. The samples differed as well, although the age of the participants was similar. Our data were collected in the [Blinded] and was 46.4% females compared the 31.25% in the Harries et al. (2005) study collected in Southern Australia. The breakdown of level of mental illness differed with the Harries et al. (2005) sample including a lower proportion of mild participants and a higher proportion of moderate and severe participants.

This study was limited by the variable SERVICES used as an indicator of potential funding in the ordinal logistic regression models. Because the [Blinded] currently uses the ICAP to determine IBA, this could affect the current services used by the individuals qualifying for HCBS waiver; i.e., there was not an independent measure of funding available. As a result, the ICAP Service Level was included in the ordinal logistic regression as a covariate and proved to be significantly involved in predicting SERVICES. However, the SIS Lifelong Learning and Health and Safety Activities subscales were still significant and moderately significant, respectively, in the ordinal logistic regression models of SERVICES. Future studies with an independent measure funding allocation are necessary to further elucidate their relation to funding.

Although the order of SERVICES from Other = 1, Supported Living = 2, Day Supported Living = 3, and Residential Habilitation = 4 made sense conceptually and allowed for clear interpretation, it failed the Score Test for Proportional Odds Assumption. As mentioned previously, the Other category was assumed to describe those who used minimal Medicaid assistance. Further study is necessary to determine if this assumption is correct and to determine where the Other category falls in this order.

## Conclusion

This study found that both the ICAP and SIS can be used for Medicaid funding ascertainment. This allows states to consider more than one instrument when deciding how to make these determinations. That the SIS is a strengths-based assessment could be an important point to consider when making the decision to use one versus the other. Indeed, our data suggest that these instruments capture distinct constructs, despite some overlap. Agencies implementing one or both of these measures should be clear about what is being measured to better align services and funding allocation to the client’s needs. Further, given the differing constructs measured, there may be instances where using both instruments, either concurrently or in serial could be beneficial, although, costs of implementing multiple instruments should be considered.

## Data Availability

The data were collected by Wyoming Institute for Disabilities to qualify for an HCBS Waiver.

## Acknowledgments

This study was funded by the University Centers for Excellence in Developmental Disabilities Education, Research, and Service Core-University of Wyoming, U.S. Department of Health and Human Services, Administration for Children and Families, Administration on Developmental Disabilities (90DDUC0011-01-00; Health Resources and Services Administration).

We would like to thank the Wyoming Department of Health for their support and collaboration on this study.

We have no conflicts of interest to disclose.

